# Epidemiological description and analysis of factors associated with cholera morbidity and mortality in Cameroon from 2018 to 2023

**DOI:** 10.1101/2024.02.20.24303118

**Authors:** Akenji Blaise Mboringong, Sen Claudine Ngomtcho, Evouna Armel, Esso Linda, Dibog Luc, Mendjime Patricia, Belinga Sandrine, Balana Flore, Theodore Ntamack, Etoundi Mballa, Roland Ndip

## Abstract

**Introduction:** Cameroon has faced frequent and severe cholera outbreaks since 1971, with high lethality. Previous studies have examined some risk factors and groups, but not for the 2018–2023 period. We analysed the profiles of cholera epidemics and determined factors associated with morbidity and mortality from cholera during this period.

**Materials and methods:** We conducted an analytical cross-sectional study using Ministry of Health cholera databases from 2018–2023. We described the socio-demographic, clinical and geographical distribution of cholera cases and used multiple regression to identify factors associated with cholera lethality.

**Results:** Between May 2018 and March 2023, there were four cholera epidemics with 18,986 cases, affecting 8/10 regions. The three coastal regions notified 83.4% (15,839/18,986) of cases. The most represented age group were those aged 25–35 years (21.9%; 4,163/1,876) and the M/F sex ratio was 1.27. The overall CFR was 2.7% (478 deaths/17,967 cases) and was highest among persons >65 years (6.8%; 59/869). Urban areas notified more cases than rural areas (13,267 vs 5,484). Factors associated with increased mortality were female sex (aOR 0.62, 95% CI 0.49-0.77), rainy season (aOR 0.60, 95% CI 0.45-0.78) and age below 45 years (aOR 0.56, 95% CI 0.45-0.69). Severe dehydration at consultation (aOR 12.76, 95% CI 7.66-21.25) was associated with higher mortality.

**Conclusion:** There were frequent cholera epidemics affecting nearly all administrative regions in Cameroon from 2018 to 2023. Factors associated with decreased mortality from cholera were female sex, living in urban areas, the rainy season, and age less than 45 years. The high caseloads and case fatality rates further reiterate the need for a multisectoral approach to the fight against cholera in Cameroon.

## INTRODUCTION

Cholera is an acute diarrheal infection caused by ingestion of food or water contaminated with the bacterium *Vibrio cholerae*. There are many serogroups of *V. cholerae*, but only two – O1 and O139, cause epidemics[1]. *Vibrio cholerae* can survive in fresh-water such as lakes [2][3] and salt-water (coastal, estuarine areas)[1]. Cholera tends to follow seasonal patterns [4][5][6] and is often associated to environmental factors such as rainfall [7] and temperature [8][9]. Some studies have clearly demonstrated a link between environmental exposure and illness in humans [10][11]. However, once introduced into the human population, interhuman transmission occurs and data suggests most of the transmission in Africa is due to direct interhuman transmission [12][13].

Since the first documented cholera pandemic in 1817, seven cholera pandemics have been recorded [14]. The seventh, that originated from South-East Asia in 1961, is still ongoing [14]. In 1970, the seventh pandemic reached the African continent, first affecting West African countries then reaching Central Africa in 1971 [15][1]. Despite having been cholera-free before 1970, Africa now bears the major burden of the global cholera cases [15]. Cholera is responsible for up to 95000 (21000-143000) fatalities worldwide every year [1].

Cameroon reported its index cholera cases in February 1971 [16]. This first epidemic led to 2167 cases according to World Health Organization reports [16]. This was followed by a 20-year period characterized by sporadic disease clusters. The cholera burden in Cameroon has increased during the past three decades, with 4026 cases in 1991, 5796 in 1996, 8005 in 2004, 10759 in 2010 and 3355 in 2014 [17][18]. However, the case-fatality rate (CFR) has decreased progressively: 12% in 1991, 8.3% in 1996, 6.3% in 2010 and 5.3% in 2014 [19][18].

Cholera epidemics have been a frequent occurrence in Cameroon especially during the past two decades with relatively high mortality and morbidity. Some previous studies have examined risk factors during earlier outbreaks [20], but not for the 2018-2023 period. Additionally, there are knowledge gaps concerning spatial and temporal distribution of recent outbreaks as well as determinants of the disease in Cameroon [20]. Filling these knowledge gaps is essential for the formulation and implementation of well-adapted cholera control strategies.

## METHODS

### Study setting

Cameroon is located in Central Africa along the Atlantic coast. It has ten administrative regions some of which, to the south are located in the equatorial rain-forest region, to the north are located in the southern part of the Sahara Desert while those in between are savanna. Cameroon has four climatic subzones [21]. The zone spanning most of the Far-North Region up to the Lake Chad Basin comprises the Sudano-Sahelian zone, the North and Adamawa Regions comprise the Tropical humid zone, most of the North-West, South-West and West Region fall within the Equatorial Monsoon zone, while the Center, South and East Regions comprise the Guinea Equatorial zone [21]. Each zone has varying lengths and time number of rainy and dry seasons with the Guinea Equatorial subzone experiencing the highest average annual rainfall (1500 to 2000mm) while the Sudano-Sahelian subzone has the shortest rainy season with an least average annual rainfall (400 to 900mm) [20].These regions are divided into 200 health districts and further into 1962 health areas [22]. Recent data suggests that the urban population in Cameroon is higher than the rural population: 58.2% versus 41.8% [23].

### Study design

This was an analytical cross-sectional study conducted at the Public Health Emergency Operations Coordination Center (PHEOCC). The study was carried out from the 1st of June to the 30^th^ of November 2023, exploiting cholera data from May 2018 to March 2023.

### Study population and sampling

The study population consisted of all confirmed cases of cholera recorded in the line-lists of the Public Health Emergency Operations Coordination Center (PHEOCC) of the Ministry of Health (MOH) from May 2018 to March 2023. The confirmed cholera cases were defined as all cases of cholera in the 2018 to 2023 cholera epidemic line-lists with documented stool culture positive for *Vibrio cholerae* serogroup O1 or cases confirmed by epidemiological link without laboratory testing.

### Database analysis

The cholera database from the PHEOCC of the MOH for the period from May 2018 to March 2023 was obtained for analysis. This database was cleaned checked for completeness, duplicity and validity of variables then analyzed for socio-demographic, clinical and epidemiological data. Seasonal data was obtained from external sources [20] and used to classify cases as having been notified during rainy or dry seasons. Population data was obtained from the MOH and used in the calculation of attack rates [24].

Epidemic curves were drawn using data on the date of onset of disease in the cases in the line lists. Attack and case-fatality rates were calculated for each epidemic, administrative region and health district using the following formulae:

**Cholera attack rate** = number of cholera cases notified during the epidemic (or in health district/administrative region)/total number of people at risk **in involved health district/administrative region**\*100,000.

**Cholera case-fatality rate (CFR) =** number of cholera deaths notified during epidemic/total number of cases notified during epidemic *100,000.

### Statistical analyses

Statistical Package for Social Science version 20 was used to analyze data. To describe geographical distribution of cases QGIS version 3.28.14 was used. Microsoft Excel 2016 for Windows was used to generate frequency tables, bar charts and epidemiological curves. Categorical variables were described using proportions and statistical analyses of proportions were done using odds ratios for comparison of variables, Kruskal–Wallis test was used to compare medians of continuous variables with asymmetrical distribution. All variables associated to cholera mortality in bivariate analysis with a significance level <0.20 were included in binary logistic multivariate analyses. The final multivariate model considered as significant, only variables with p <0.05. During bivariate and multivariate analyses, missing data was handled using the pairwise deletion approach.

### Ethical and administrative considerations

Authorization to use MOH data was obtained from the PHEOCC in Yaounde, Cameroon. Ethical clearance was obtained from the University of Buea Faculty of Health Sciences Institutional Review Board. This analysis was done on a codified secondary dataset so it was not possible to involve the patients or the public in the designing, execution, or dissemination of the results of the study.

## RESULTS

### Distribution of cases by administrative region and health district

Between 2018 and 2023, Cameroon experienced four cholera outbreaks. The first affected the North, Center and Far-North Regions with a total of 1,628 cases from 2018 to 2019. This epidemic affected 34 health districts (HD): 08 in the Far North, 13 in the North Region and 13 in the Center region. The epicenters of this epidemic were Garoua I (309 cases), Pitoa (213 cases) and Bibemi (209 cases) districts.

The second was confined mainly to the Bakassi health district (359 cases) and the Ekondo-Titi health district (1 case) in the South-West Region. It lasted two months-November to December 2019.

The third outbreak began in the Littoral Region in January 2020 and spread to the Center, South and South-West Regions, resulting in 1,889 cases by July 2020 when it ended. It encompassed epidemic affected 18 health districts: 10 in the Littoral: Cite, 02 in the South-West Region, 03 in the Center region and 03 in the South Region. The most affected health districts were Kribi (762 cases), Nylon (299 cases) and Bonassama (214 cases).

The fourth began on October 15th 2021, in the South-West Region and by March 2023, eight regions had reported cases, with the majority of them occurring in the Littoral (7,412 cases) and South-West (6,027 cases) Regions. This prolonged outbreak affected 61 health districts: 18 in the Center Region, 06 in the Far North, 01 in the North Region, 02 in the East Region, 18 in the Littoral, 04 in the South Region and 12 South-West Region The distribution of cases by administrative region and health district is illustrated in **Fig 1 (Supplementary material S1).** The most affected districts were Bangue (485 cases), Bakassi (309 cases), Bonassama (645 cases), Buea (935 cases), Cité des Palmiers (707 cases), Deido (1,997 cases), Djoungolo (337), Ekondo-Titi (353 cases) and Limbe (3,136 cases).

**Fig 1:**
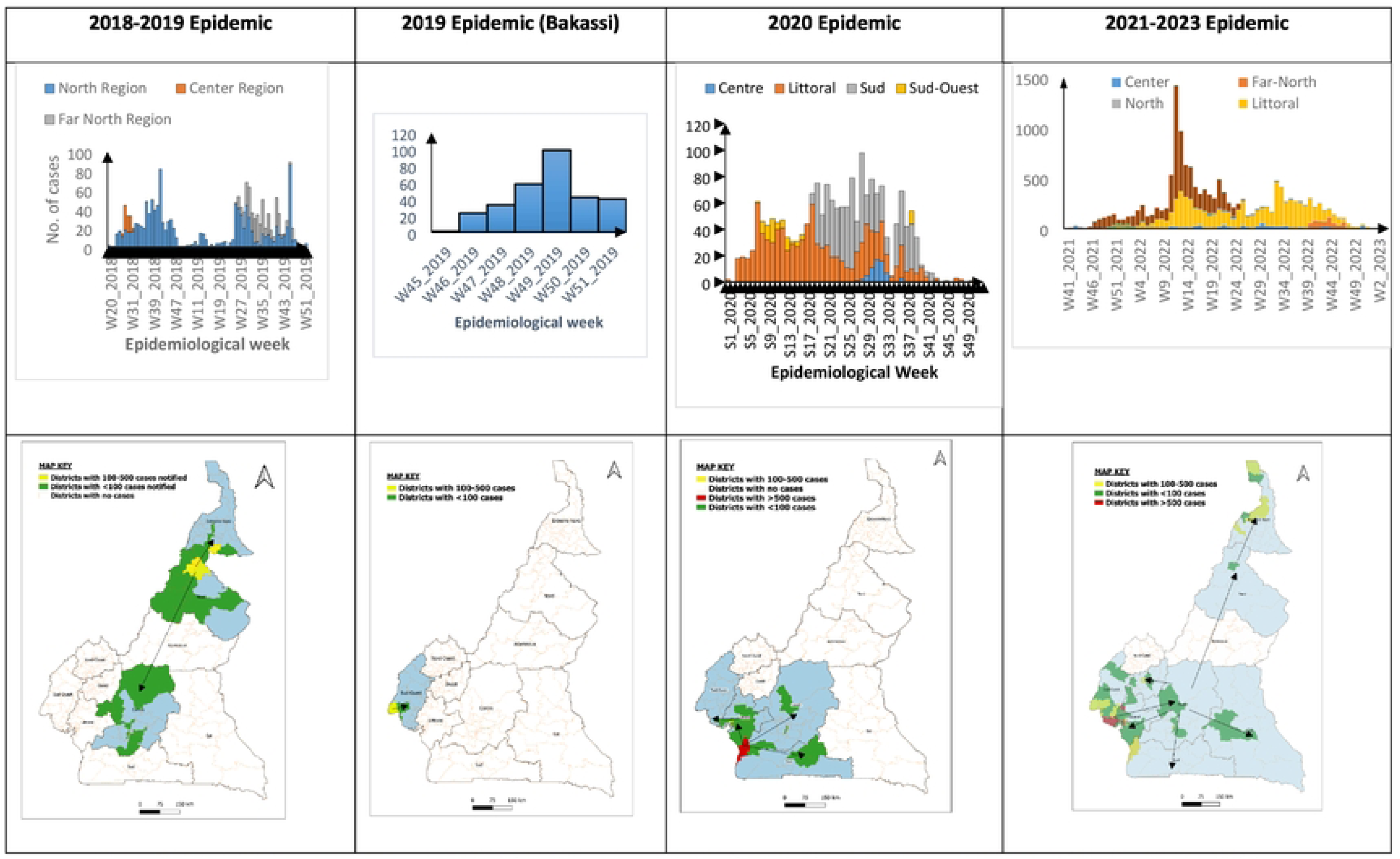
Epidemic curves and geographical distribution of cases by health district of cholera cases during each epidemic from 2018-2023. The maps in the second row indicate the districts affected during the corresponding epidemics indicated in the epidemic curves in the row above. The arrows in the maps indicate the evolution of the spread of cases by region /districts affected. This information was generated from the dates of onset of cases notified in the cholera database.

The coastal Regions (Littoral, South-West, South) accounted for 83.4% (15,839) of the 18,986 cases reported during the study period (**Table 1)**. A total of 85/200 (42.5%) of health districts in Cameroon were affected by cholera between May 2018 and March 2023.

**Table 1:** Number of epidemics, their duration and number of cases notified in Cameroon from May 2018 to March 2023.

| **Epidemic** | **Region** | **Date of onset** | **Date of last notification** | **No. of cases** | **Total no. of cases** |
| --- | --- | --- | --- | --- | --- |
| **2018-2019** | North | 18/05/2018 | 17/12/2019 | 1,212 | **1,627** |
|  | Center | 11/07/2018 | 12/08/2018 | 66 |  |
|  | Far North | 01/07/2019 | 23/11/2019 | 350 |  |
| **2019** | South-West<br>(Bakassi HD only) | 13/11/2019 | 23/12/2019 | 359 | **359** |
| **2020** | Center | 13/07/2020 | 05/08/2020 | 62 | **1,889** |
|  | Littoral | 04/01/2020 | 18/12/2020 | 967 |  |
|  | South | 01/05/2020 | 18/10/2020 | 780 |  |
|  | South-West | 22/02/2020 | 19/09/2020 | 80 |  |
| **2021-2023** | South-West | 15/10/2021 | 1/9/2022 | 6,027 | **15,111** |
|  | Littoral | 18/11/2021 | 29/12/2022 | 7,412 |  |
|  | Center | 23/10/2021 | On-going | 707 |  |
|  | Far North | 01/01/2022 | 15/12/2022 | 486 |  |
|  | North | 14/02/2022 | 16/6/2022 | 50 |  |
|  | South | 10/12/2021 | 15/5/2022 | 214 |  |
|  | West | 14/04/2022 | 3/12/2022 | 203 |  |
|  | East | 13/07/2022 | On-going | 12 |  |
| **TOTAL** |  |  |  | **18,986** | **18,986** |

### Age and Sex distribution of cases

The most represented age group was the 25 to 35-year age group (22.2%; 4,136 cases), followed by the 15 to 25-year age group (21.4%; 4,012 cases) and the 35 to 45-year age group (14.7%). The 0 to 5-year age represented 9.5% of the total caseload. The overall male: female sex ratio was 1.27 (10,594 males/8,334 females).

### Attack rates and case-fatality rates

The 2019 epidemic in the South-West region (Bakassi) had the highest attack rate (388.5/100,000 inhabitants at risk) followed by the on-going 2021-2023 epidemic (170.8/100,000 inhabitants at risk). The coastal regions (Littoral, South-West and South) recorded higher attack rates compared to the rest of the regions as shown in **Table 2**. The Center Region despite being frequently in epidemic had the lowest attack rate (14.9/100,000). The coastal regions had the most cases but conversely, had the lowest death rates (Littoral: 2.7%; South: 2.8%; South-West: 1.7%). The overall CFR was 2.7%. The CFR increased with age. It was highest among cases over 65 years old (6.8%) and lowest for cases aged 15-25 years (1.3%). The CFR decreased with each passing (epidemic 4.9% in the 2018-2019, 4.4% in the 2019 (Bakassi outbreak), 4.2% in the 2020 and 2.1% in the 2021-2023 outbreaks.

**Table 2:** Attack rates of cholera epidemics per region from 2018 to 2023 in Cameroon.

| **Region** | **Total cases notified** | **Total population at risk** | **Attack rate/100000** | **Total Alive** | **Total Dead** | **CFR (%)** |
| --- | --- | --- | --- | --- | --- | --- |
| **Center** | 813 | 9,037,324 | 9.0 | 695 | 22 | 3.1 |
| **East** | 12 | 44,683 | 26.9 | 10 | 2 | 16.7 |
| **Far North** | 832 | 1,146,878 | 72.5 | 653 | 35 | 5.1 |
| **Littoral** | 8,328 | 7,502,297 | 111 | 7,505 | 211 | 2.7 |
| **North** | 1,262 | 267,803 | 47 | 1,182 | 64 | 5.1 |
| **South** | 993 | 480,552 | 206 | 923 | 27 | 2.8 |
| **South-West** | 7,166 | 1,737,783 | 412 | 6,357 | 110 | 1.7 |
| **West** | 203 | 775,848 | 27 | 164 | 7 | 4.1 |
| **Total** | 19,609 | 23,396,168 | 83.8 | 17,489 | 478 | 2.7 |

### Severity of dehydration at time of consultation

Twenty percent (3,311/16,550) of cases had mild dehydration, 49% (811/16,550) had moderate dehydration and 31% (5,123/16,550) had severe dehydration at time of consultation. The proportion of cases with severe dehydration was highest among those older than 65 years (39.1%; 299/764) and lowest among the cases aged 0-5 years (21%; 319/1,519). The median ages of cases with mild, moderate, and severe dehydration were 25.0 years, 27.0 years, and 29.0 years, respectively. The Kruskal-Wallis test was used to compare these medians and indicated a significant association between increasing age and an increase in the severity of dehydration (p-value<0.0001).

### Seasonal variation of cholera cases

Apart from the Sudano-Sahelian zone which experienced higher case-notification in the dry seasons, all the other sub-climatic zones experienced higher case notification during the rainy seasons. Peaks occurred during the rainy and dry seasons but overall, more cases were notified during the rainy seasons as shown in **Fig 2 (Supplementary material S2)**.

**Fig 2:**
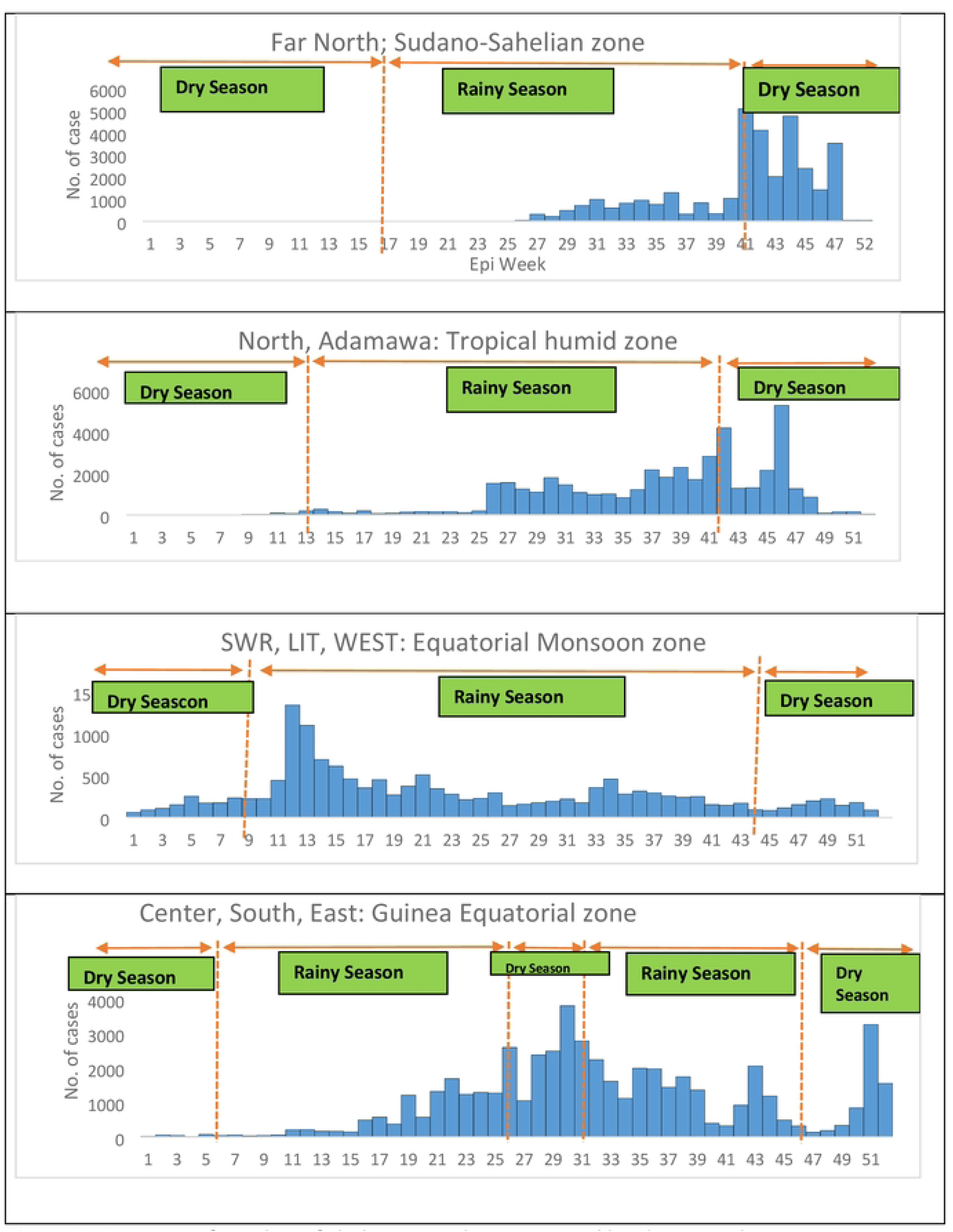
Variation of number of cholera cases by season and by climatic subzone in Cameroon, 2018-2023. This classification of climatic subzones and the periods of dry and rainy seasons were obtained from the scientific publication by Cornelius et al [21].

### Factors associated with mortality

On bivariate analysis, there were significantly higher odds of death among cases notified in rural health areas than among those notified in urban health areas (OR 1.68, CI: 1.38-2.03) and significantly higher odds of death among male cases (OR 1.97, CI: 1.70-2.28). We found significantly higher odds of death among cases older than 35 years, with the highest odds in persons older than 65 years (OR 4.29, CI: 2.17-6.77). Severe dehydration, (OR 13.7, 8.28-22.70) also appeared to be significantly associated with increased cholera mortality. However, multivariate regression analysis revealed that of these, only age greater than 45 years and severe dehydration were predictors of increased cholera mortality while the dry season and female sex appeared to protect against dying from cholera **(Table 3).**

**Table 3:**
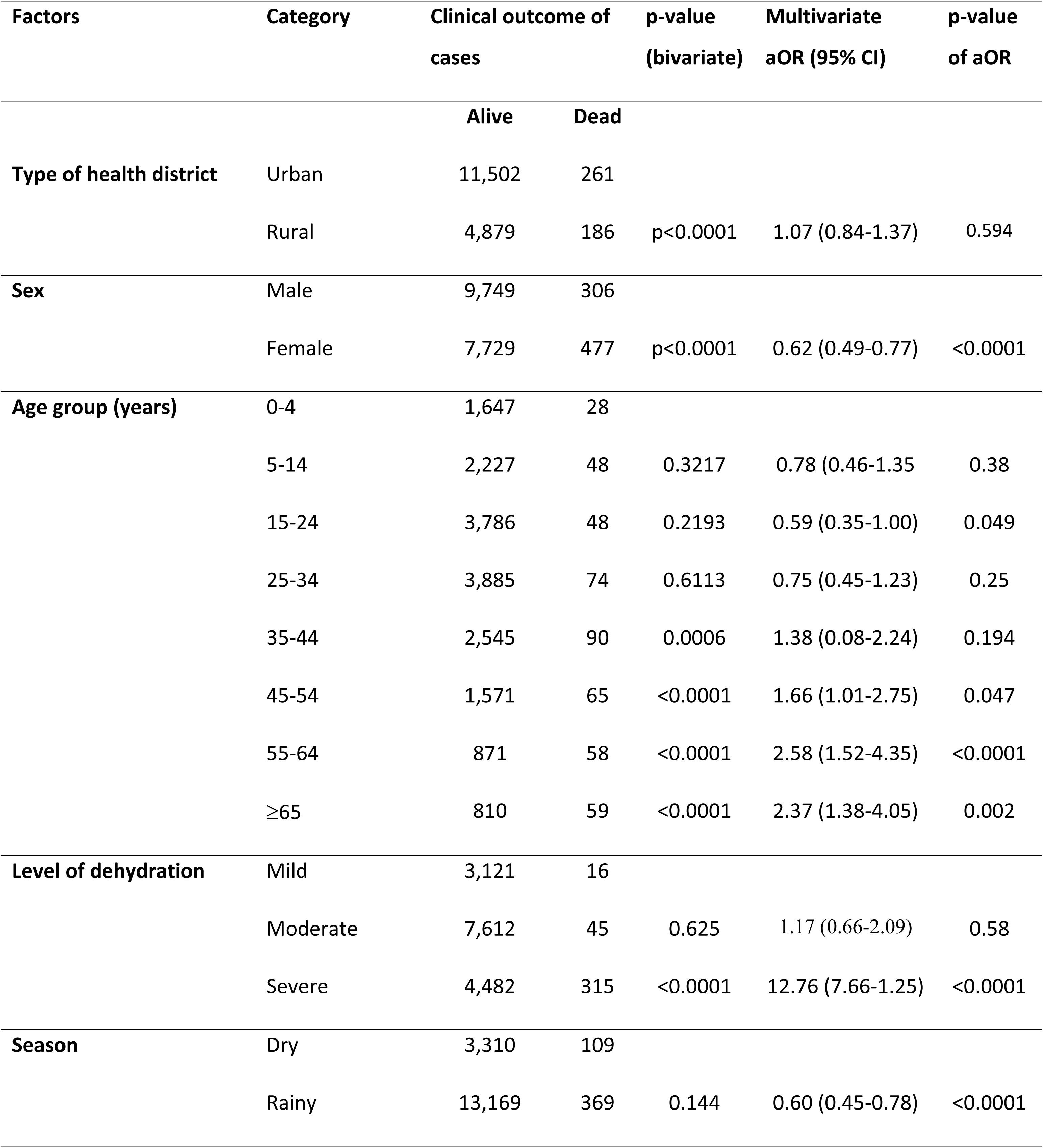
Bivariate and multivariate analyses of factors associated with cholera mortality in Cameroon, 2018-2023.

## DISCUSSION

The findings in this study have provided an insight to the epidemiological profiles of cholera epidemics that have occurred in Cameroon from May 2018 to March 2023.

During the study period, there were four cholera epidemics, and all regions except the Adamawa and North-West Regions, were affected. Of the cases, the North and Far North Regions reputable for being frequently affected by cholera epidemics accounted for only 11.1% of cases within this period while the South-West, Littoral and South Regions (the coastal regions) accounted for 84% of cases.

Compared to the 2004-2013 cholera situation in Cameroon, our study revealed a decrease in the proportion of cases notified by the two northern regions, which during that period accounted for 47.3% of the cases [25]. The Littoral Region, the South-West Region and the South Region from 2004-2013 notified 38.7% of cases [25]. Despite this shift in distribution of cases, similar to the cholera distribution from 2004-2013, the north regions and the three coastal regions accounted for the majority of cases put together. This was not surprising because recent data show that these regions are major cholera hotspots [26]. Several authors have demonstrated a higher susceptibility of coastal areas to cholera epidemics associated to the survival of *Vibrio cholerae* in marine organisms such as copepods which are zooplankton [27][28].The history and scale of cholera pandemics indicate that coastal areas are the main sources of cholera epidemics, which then spread to inland regions through other routes. Coastal ecosystems have a high abundance of plankton, which are closely linked to cholera bacteria [29]. This, compounded by the overcrowding and inaccessibility to potable water in cities like Douala in the Littoral Region and cross-border movements between South-East Nigeria and the South-West Region of Cameroon due to political unrest could explain the high susceptibility of these Regions to cholera [30]. The frequent cholera epidemics in the northern regions of Cameroon have been linked to socio-cultural, economic and climatic factors particularly inaccessibility to clean water due to frequent droughts, mistrust in public health messages (due to multiplicity of ethnic languages, dependency and sentimentality toward animal husbandry), cultural practices of eating food from one plate, hospital avoidance (due to belief that traditional healers cure cholera)[31].

In this study, urban health areas notified more than twice the number of cases notified in rural health areas (13267 versus 5484 cases), that is, 70.8% of the caseload. The urban population in Cameroon is greater than the rural population: 58.2% versus 41.8% [23], but the disparity in the proportion of cases reported was still high (70.8% vs 29.2%). A previous systematic review of the cholera situation in sub-Saharan Africa had already revealed that a large proportion of the cholera burden occurs in urban areas, particularly in dense urban settings (Zerbo *et al.*, 2020). No previous study in Cameroon has evaluated the susceptibility of rural versus urban areas to cholera but the results were somewhat unexpected as a recent WHO publication indicates there is better access to basic drinking water and sanitation services in urban areas compared to rural areas in Cameroon [33]. What may account for the high number of cases though, may also be the fact that the urban areas notifying most of our cases were also coastal towns (Kribi, Douala, Limbe, Tiko).

During our study period, there were considerably more male cases than female cases (male: female ratio of 1.27). According to World Bank population data, females made up 50.1% of the Cameroonian population between 2018 and 2023 so despite there being more females than males, overall males appeared more susceptible than females to cholera [34]. The findings are similar that of the 2011 to 2013 cholera epidemic study in Haiti [35] and in Sierra Leone [36]. An analysis of the cholera situation in Nigeria, a neighboring country which is also frequently in epidemic also revealed a male-predominance in terms of incidence of cholera cases but there is not enough evidence to attribute the disparity in rates in which males and females are affected to physiological causes. Rather, socio-cultural (gender) roles played by males and females may be able to explain these disparities [37]. The median age of cases was 27 years with ages from 15 to 35 years being the most represented. Previous studies in Cameroon have revealed similar findings. For example, Nsagha and collaborators found that in Buea, the average age of cholera cases during the 2010-2011 epidemic was 29 years [38]. Similarly, WHO reports show that the 2022 cholera epidemic in Malawi affected mostly the 21-30 years age group [39]. In areas endemic with cholera, younger children (less than five years of age) are more susceptible to having cholera while in non-endemic areas adults tend to be more affected [40]. Our findings align with this as cholera is not endemic in Cameroon.

Attack rates varied widely between regions, with the Littoral, South and South-West Regions having the highest attack rates. These were also the regions that notified the highest number of cases. During the 2004-2013 period, the Littoral and South-West regions also recorded the highest attack rates together with the two northern regions [25]. Most of the cases recorded in the Littoral Region were in the city of Douala, a densely populated coastal city, which could be the reason behind the high-attack rate in the Littoral Region. The case-fatality rates (CFRs), however, showed an inverted pattern to the attack rates in these regions as these were the three regions with the lowest case-fatality rates of all ten regions. Many of the cases in these regions were notified in urban health districts in which there is better access to healthcare, i.e., quicker notification, detection and treatment. A study in 2016 in the Far North region revealed that most cholera deaths occurred outside of health facilities [41]. The reasons for the high rates of death out of the health facilities were not evaluated in our study but according to Djouma and collaborators, they were due to delays in deciding to go to a hospital and the preference to seek treatment in community treatment centers rather than hospital-based treatment centers [41]. Mistrust for public health messages is high in these Regions and may have contributed to the high mortality from cholera [31].

The case-fatality increased with increasing age, with the age group above 65 years having the highest CFR (6.8%). This finding was supported by an extensive literature review by the Global Task Force on Cholera Control which revealed multiple studies had found increasing age to be associated with higher CFRs especially as from ages greater than 40 years [42]. The recent cholera epidemic in Malawi also has similar demographics with people older than 60 years having the highest CFR [39]. The CFR among children less than 5 years of age during this period was low despite this group being particularly vulnerable to cholera.

Using binary logistic regression, male sex, age above 45 years and the dry season were predictors of increased mortality from cholera. Children are generally more prone to death from cholera infection due to difficulty in the clinical management (e.g. fluid management) of pediatric cholera cases as well as their higher susceptibility to malnutrition which exacerbates the risk of cholera-related death, especially in children aged less than five years. However, in our study, people who were 45 years and older had higher chances of dying from the disease. Many recent epidemics in sub-Saharan Africa have had similar patterns [43] [37] [42]. Dalhat and collaborators, in Nigeria suggested that some possible reasons for this increased mortality among the older population could be lack of proper care, dependence on family members for support or presence of other health conditions and undernutrition [43]. However, some other studies still identify younger ages as having higher risk of dying from cholera [38]. In our study, severity of dehydration was associated with increasing age (Kruskal-Wallis p-value < 0.0001). These findings may be explained by the reluctance or difficulty in displacement of older people to seek care in treatment centers. This is a hypothesis that has to be further evaluated as it may significantly reorientate the public health response during cholera epidemics.

The higher rate of male deaths from cholera has also been cited by several studies [37]. In a systematic review of 78 studies seventeen, studies compared the sex-specific CFRs using statistical models or tests. Of these, nine found no statistical difference, seven reported a greater CFR in men and one reported more female deaths. Another 21 studies provided sex-specific CFRs (or data to compute them) but they did not compare them with tests. In 12 studies, CFR was higher in males. Half of the included studies (39/78) did not report CFR for males and females [42]. However, the higher lethality in males has not been attributed to physiological differences in sex but rather to the societal roles played by the different genders in communities affected by cholera [44].

Irrespective of zone-specific climatic conditions, more cases were reported in the rainy season than in the dry season. Cholera data analysis in Cameroon from 2004-2012 is supportive of these findings as it reveals that cholera cases surged mostly during the rainy seasons and at the end of the dry seasons [25]. A kenyan study showed that depending on the time of the year, decreased rainfall could lead to more cases but also, increased rainfall leading to floods also led to an increase in cholera cases [45]. On multivariate analysis, the rainy season appeared be associated with lower mortality (aOR 0.60, 95% CI 0.45-0.78). A modelling study in Iran showed that low precipitation combined with high temperatures (as observed during the dry season) were associated with an uptick in cholera cases [46]. The possible reason for this could be that *Vibrio cholerae* replicates faster under these conditions [46]. However, despite our study and several other studies reporting an increase of cholera cases in the rainy season [20][47], no studies evaluating association between mortality from cholera and seasonal changes were found during literature review.

Rural areas were not associated with higher rates of death than urban areas (aOR 1.07 95% CI 0.84-1.37). Though few, some studies comparing cholera death rates in rural and urban areas have been published indicating higher mortality in rural than in urban areas [48][35][37]. Reasons proposed in other studies for the higher mortality in rural areas include delayed access to care either because of hesitancy or insufficient number of cholera treatment centers[41][37]. However, the database we analyzed did not permit us to directly impute this factor.

## STUDY LIMITATIONS

This study had a few limitations which did not very much impact the quality of the results but that are worth mentioning. Firstly, secondary data (data collected for other purposes) was used for the analysis of the epidemiological profiles, meaning, there was a considerable amount of missing data for several variables. However, the very large sample size from ensured that the study still had enough statistical power. Pairwise deletion was attempted in the multivariate analysis, but led to inappropriate models so all cases with missing data were deleted. Binary logistic regression analysis was conducted with a smaller subset (about 80 percent) of the initial database. Finally, the data did not also permit us to evaluate some important factors of cholera death and severity such as time lapse between onset of symptoms and consultation, and also place of death of cases (community or health facility).

## CONCLUSIONS

Between 2018 and 2023 there were four distinct cholera epidemics in Cameroon. All but two regions (North-West and Adamawa) notified cases. Urban health areas notified roughly twice the number of cases than rural health areas. Factors associated with increased cholera mortality were male sex, age above 45 years and severe dehydration at time of consultation. Despite the majority of cases occurring during the rainy seasons, mortality was higher in the dry seasons.

## RECOMMENDATIONS

Due to the high frequency of epidemics and large number of cases of cholera, we recommend that the Ministry of Public Health should improve sensitization of the population to in order to increase population awareness on preventive measures. Also, a multisectoral approach to cholera response in the country will be beneficial in terms of reducing caseloads and deaths from cholera.

High-risk groups for cholera mortality (male sex and age above 45 years) were identified but the study design did not permit identification of the reasons behind these findings. It is imperative to conduct further studies on these, using designs better suited to provide explanations to these findings and to overcome the limitations of this study.

## Data Availability

Data are already included in the tables and figures in the manuscript.

## SUPPORTING INFORMATION

**S1 Fig:** Fig 1: Epidemic curves and geographical distribution of cases by health district of cholera cases during each epidemic from 2018-2023. The maps in the second row indicate the districts affected during the corresponding epidemics indicated in the epidemic curves in the row above. The arrows in the maps indicate the evolution of the spread of cases by region /districts affected. This information was generated from the dates of onset of cases notified in the cholera database.

**S2 Fig:** Fig 2: Variation of number of cholera cases by season and by climatic subzone in Cameroon, 2018-2023. This classification of climatic subzones and the periods of dry and rainy seasons were obtained from the scientific publication by Cornelius *et al*. [21].

## Notes

### Competing Interest Statement

The authors have declared no competing interest.

### Funding Statement

This work was not funded by any grant donor or partner. The authors were responsible for the financing of the entire research project and the publication process.

### Author Declarations

The Institutional Review Board of the University of Buea, Cameroon.

